# Trends in Efficacy, Safety, and Tolerability of Approved Multiple Sclerosis Disease Modifying Treatments

**DOI:** 10.1101/2023.12.11.23299815

**Authors:** Kyle Valker, Kangho Suh

## Abstract

**Background:** Disease-modifying therapies (DMTs) have become the mainstay of treatment for relapsing forms of multiple sclerosis (MS), reducing relapse rates and slowing disease progression.

**Objectives:** To determine whether or not available MS DMTs have demonstrated an increase in safety, efficacy, and tolerability over time.

**Methods:** Results from pivotal phase III trials of approved MS DMTs were used to create a dataset of relevant outcomes. Common endpoints analyzed include annualized relapse rates (ARR), rates of serious adverse events (SAE), and rates of discontinuation due to adverse events. Trial comparator, active or placebo, was also documented. Descriptive statistics and Fisher exact tests were performed on outcomes stratified by recency of pivotal trials.

**Results:** On visual inspection, there was a trend of decrease in ARR. A significant relationship was seen between recent approvals and trial design with an active comparator (p=0.004), as well as between recent approvals and ARR (p=0.020). No significance was found between recent approvals and SAE (p=0.138), formulation and discontinuation (p=0.559), or recent approvals and formulation (p=0.352).

**Conclusion:** DMTs for relapsing forms of MS increased in efficacy over time. Oral therapies offered similar tolerability to other routes of administration. Further research is warranted to identify if these clinical trial findings translate to real world evidence.

**What was already known:**

- The number of FDA approved disease-modifying therapies for multiple sclerosis has been steadily increasing. Available routes of administration include injectable, oral, and infusions.
- These medications are proven to be effective in reducing MS relapse rates and slowing overall disease progression, with varying degrees of safety and tolerability.
- The comparative efficacy of these therapies varies, with certain medications often deemed high efficacy.

**What this study adds:**

- We used published phase III trial results for each medication to provide a direct comparison between each medication’s efficacy, safety, and tolerability at time of approval.
- Our analysis demonstrates a trend in increasing efficacy of available therapies along with the use of active comparators for controls in disease modifying treatments for multiple sclerosis.

## Background

Multiple sclerosis is a chronic autoimmune central nervous system (CNS) disorder characterized by neuronal demyelination. Prevalence of the disease varies by region, seen significantly more commonly in North America and Europe.^1^ Disease course varies by individual, often broken down into four subtypes: the majority of patients present with a relapsing remitting course (RRMS) at disease onset, characterized by acute symptomatic exacerbations with a full or nearly complete return to function afterwards.^2^ The most widely used diagnostic criteria for the disease is the McDonald criteria^3^ which combines clinical presentation and history, lesions found on MRI results, and most recently the presence of CSF-specific oligoclonal bands. Clinical presentation of the disease is complex, with a wide range of potentially debilitating symptoms manifesting particularly during acute relapses, requiring a multifaceted approach to treatment.

The mainstay of multiple sclerosis therapy has become disease-modifying therapies (DMTs) since their emergence in the 90s. These therapies exert immunomodulatory effects and have demonstrated reductions in the risk of relapses and delaying disease progression.^4^ Approved indications vary by drug however every therapy currently available is approved for treatment of relapsing-remitting multiple sclerosis. DMTs also vary by route of administration: injection, oral, or infusion. Newer therapies, particularly monoclonal antibodies which exert more direct immunomodulatory effects, are generally thought to be the higher efficacy approach to treatment.^5^

### Objective

The primary objective of this study was to summarize trends in reported efficacy and safety data for every currently approved DMT, examining whether or not newer agents have demonstrated equal or greater efficacy and improved safety and tolerability outcomes.

### Methods

PubMed was utilized to collect pivotal phase III trials for all currently FDA approved DMTs, with trials dating between 1993 through 2022.^6-29^ Generic equivalents, novel formulations of existing medications, and monomethyl fumarate, an active metabolite of dimethyl fumarate approved based on bioavailability data, were excluded. Safety and efficacy data common amongst each trial was pulled to create a dataset. Therapies were also categorized based on route of administration: injectable, oral, or infusion. The primary efficacy endpoint that was identified and collected from every trial was annualized relapse rate (ARR). Other endpoints included in the dataset when available were percentages of patients with serious adverse events as a measure of safety and percentages of patients with adverse events leading to discontinuation as a measure of tolerability. Trial comparator, active or placebo, was also collected.

ARR was grouped by route of administration and plotted against time to visualize potential trends. Based on our objective to assess efficacy and safety in recently approved DMTs compared to older DMTs, we chose *a priori* to run several Fisher’s exact tests for recently approved DMTs in the past 10 years (1993-2012 or 2013-2022) by 1) pivotal trial-reported ARR (<0.2 or >0.2), 2) annual serious adverse effect (SAE) probability (< 5% or ≥5%), 3) formulation (oral vs injection/infusion), and 4) trial comparator, active or placebo. Additionally, we assessed whether there was a relationship between tolerability (i.e., annual discontinuation probability <5% vs ≥5% due to adverse event) and formulation. Level of significance was set at p = 0.05 for all tests. ARR was deemed the primary measure of overall efficacy as it was reported in all trials gathered and the primary efficacy endpoint in the vast majority. A cutoff of 0.2 was chosen based on the mean and median of reported ARRs since a point of clinical relevance was unclear. Annual probability of SAE was used as the primary measure of safety. Annual probabilities of SAE and discontinuation due to AE were calculated from respective percentages of patients from each trial. Due to limitations in available data, safety and tolerability outcomes were assessed over the last 12 years of the dataset (Table 1), resulting in cutoffs of 2009-2016 or 2017-2022.

**Table 1.**
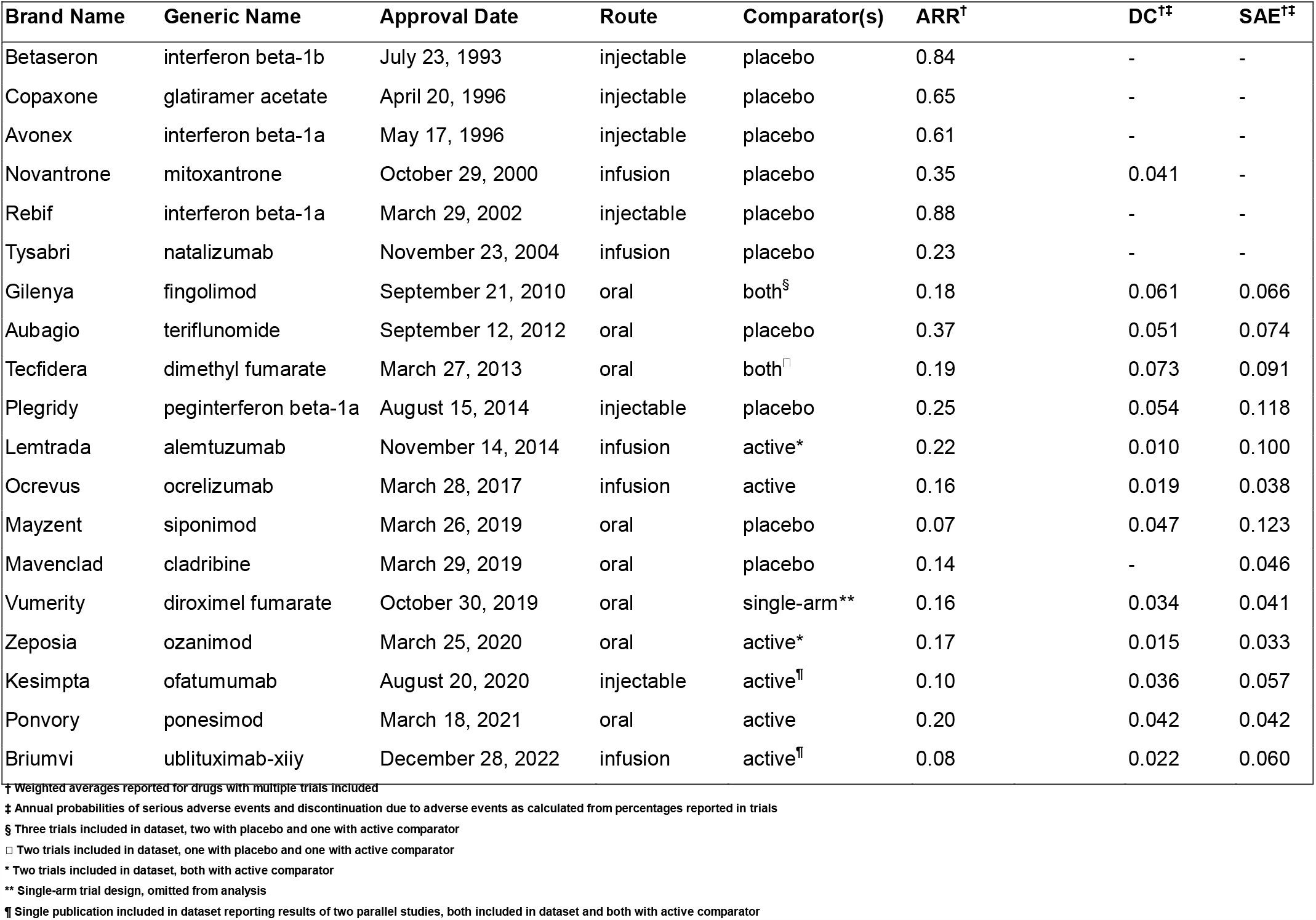
Characteristics of multiple sclerosis disease modifying treatments.

## Results

A total of 25 pivotal phase III trials were included in the dataset. All were randomized controlled trials with the exception of the single-arm EVOLVE-MS-1 trial.^26^ DMTs, regardless of treatment modality, showed a decline in ARR over the 30-year period demonstrating a general trend in increased efficacy (Figure 1). This was supported by a significant association between ARR less than 0.2 and more recent DMT approvals (p=0.020). However, for SAE categorized at above and below 5%, we did not find a significant relationship with recently approved drugs (p=0.138).

**Figure 1.**
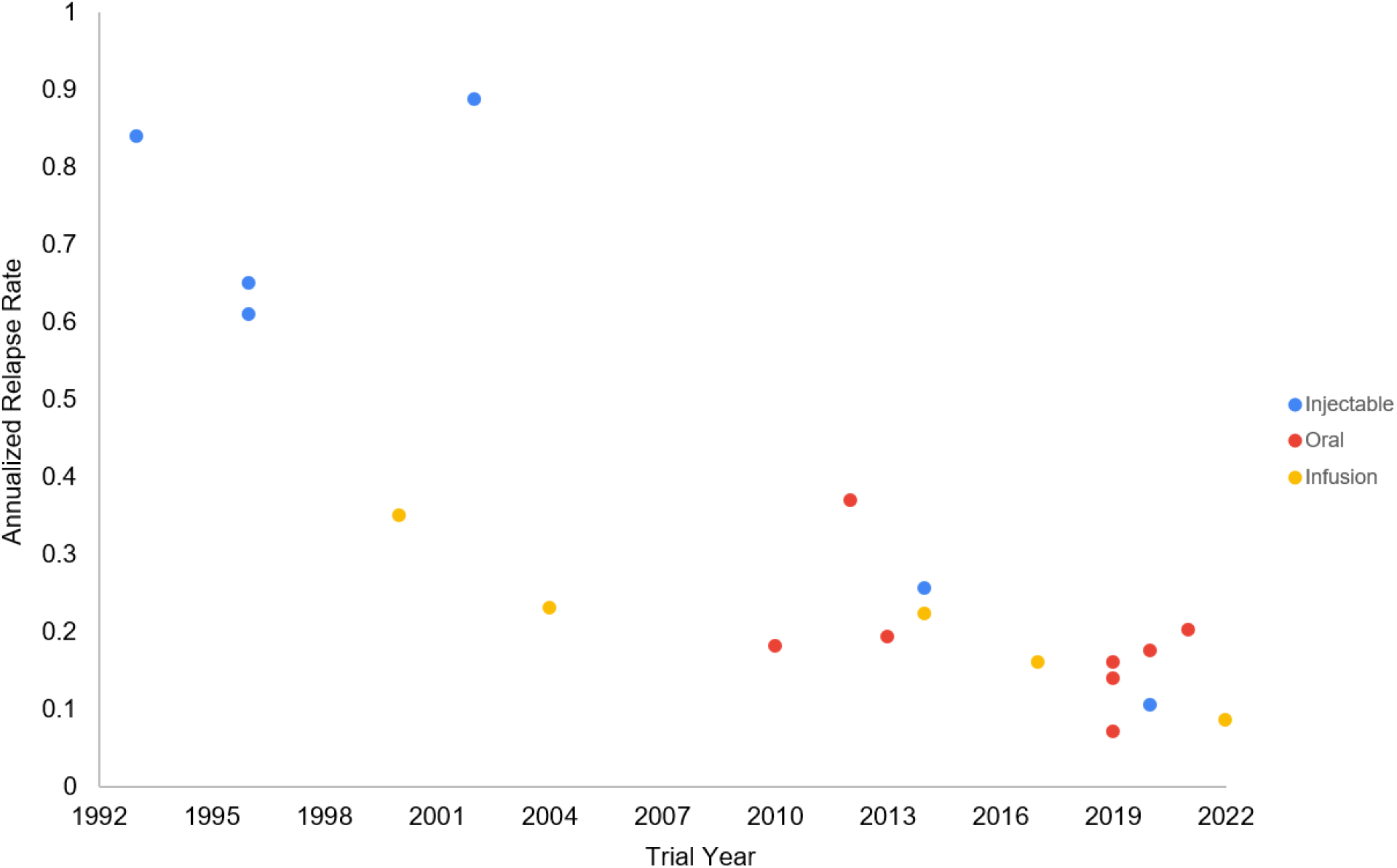
Trends of annualize relapse rate by formulation type.

The other characteristics that we decided to investigate were related to mode of administration (oral vs. injection/infusion). Tolerability was assessed at a cutoff of 5% annual discontinuation probability, and there was no significant association with oral vs. injection/infusion (p=0.559). Additionally, while there seemed to be a shift in developing more oral formulations as a higher proportion of recently approved DMTs were oral (n=6/11) compared to older approvals (n=2/8), this was not statistically significant (p=0.352). We did find recently approved DMTs had a positive significant association in the use of an active comparator vs. placebo (p=0.004).

## Discussion

To our knowledge, this is the first study examining trends in efficacy, safety, and tolerability of MS DMTs inclusive of all currently approved therapies. Our findings support the notion that newer therapies are generally more effective. Should this trend continue MS will potentially become increasingly more manageable. Although no currently approved therapy is truly curative, the increasing potential to reduce probability of acute relapses, and by extension overall disease progression, offers hope for a future where patients with MS may experience a far lower average disease burden than historically.^30^ The lack of a trend towards increasing safety is noteworthy; these therapies still undoubtedly have a positive risk/benefit profile for the vast majority of patients, however a greater focus on safety outcomes could be an area of focus in developing future therapies. Notably the trials referenced for our database focused specifically on RRMS, and indeed the majority of DMTs available are approved for management of RRMS and SPMS. Currently only ocrelizumab is FDA approved for PPMS and only mitoxantrone for PRMS. Other therapies may be used off-label for PPMS and PRMS, however their comparative efficacy and safety profiles are less clear in these contexts.^31^ These subtypes warrant further study to identify optimal medication therapy management and potential novel treatments.

Our findings on tolerability by route of administration were interesting given the widespread reputations of certain therapies included. Interferons, for instance, are notorious for significant dose limiting flu-like symptoms.^32^ Injections and infusions also tend to have very high rates of injection site/infusion associated reactions. Notably the use of probability of discontinuation due to AEs may not fully elucidate tolerability of each drug as factors outside of AEs also weigh in. For example, injection anxiety^33^ is an otherwise noted barrier to access for many individuals when choosing a prospective DMT; many patients may find frequent injections to be intolerable and avoid such medications altogether. With this in mind, the trend towards development and approval of comparably safe and effective oral therapies is altogether sensible, offering a more suitable option for many patients. Unfortunately many currently approved oral therapies also have a number of potentially dose-limiting AEs^34^, such as gastrointestinal upset, which is likely why our results indicated no significant difference in tolerability between oral therapies and alternatives when examining impact of AEs in isolation. Managing adverse effects of DMTs and increasing tolerability overall going forwards is an important consideration given the impact adherence to DMT regimens is noted to have on improving patient outcomes and reducing costs.^35^

The clear trend towards trial designs featuring active comparators in our dataset was to be expected given ethical considerations of withholding treatments that have increasingly become widely available, affordable, and proven effective in reducing burden of disease. Notably other studies have also acknowledged a shift towards implementation of active comparators in clinical trials.^36^

This study has several limitations. Generally, earlier trials were more limited in terms of reported outcomes, particularly in regard to MRI data and specific safety outcomes, limiting what could be included. Selected outcomes for this study were chosen on the basis that they were reported amongst all trials included over the respective date ranges chosen. However, we selected what we believed to be most RRMS such as ARR. Trial duration also varied, ranging from 48 to 104 weeks. We adjusted for this by using annualized rates. Another limitation was that earlier trials, particularly those conducted before widespread implementation of the McDonald Criteria as a standard for diagnosis, likely enrolled participants with a higher baseline disease burden when compared to contemporary trials. In addition, certain outcome measures such as measures of disability or severity of adverse effects have an inherent degree of subjectivity by nature, further complicated once again by differences in protocols between trials in defining such outcomes. The consolidation of IM injectables and IV infusions into one subgroup for analysis may also have implications as outcomes of interest may vary between these different routes of administration. However, for the purpose of comparing oral therapies to currently available alternatives, this grouping does provide clear insight.

## Conclusion

The number of FDA approved MS DMTs has gradually increased over the past 30 years. Data from phase III trials pivotal to each drug’s approval supports trends in increasing efficacy but not safety over the period. Despite increasing adoption, there was no demonstrable advantage of oral therapies over injections/infusions in terms of tolerability, and there has not been a significant shift towards oral therapy approvals. Studies have shifted towards inclusion of an active comparator rather than a placebo over time. Reported results are limited to patients with a diagnosis of RRMS; other subtypes of MS warrant further study.

## Data Availability

All data produced in the present work are contained in the manuscript

## References

1. Leray E, Moreau T, Fromont A, et al. Epidemiology of multiple sclerosis. Revue Neurologique 2016;172(1):3–3. doi: 10.1016/j.neurol.2015.10.006

2. Rejdak K, Jackson S, Giovannoni G. Multiple sclerosis: a practical overview for clinicians. British Medical Bulletin 2010;95(1):79–79. doi: 10.1093/bmb/ldq017

3. Thompson AJ, Banwell BL, Barkhof F, et al. Diagnosis of multiple sclerosis: 2017 revisions of the McDonald criteria. The Lancet Neurology 2018;17(2):162–162. doi: 10.1016/s1474-4422(17)30470-2

4. De Angelis F, John NA, Brownlee WJ. Disease-modifying therapies for multiple sclerosis. Bmj 2018;363:k4674. doi: 10.1136/bmj.k4674 [published Online First: 20181127]

5. Samjoo Imtiaz A, Worthington E, Drudge C, et al. Efficacy classification of modern therapies in multiple sclerosis. Journal of Comparative Effectiveness Research 2021;10(6):495–495. doi: 10.2217/cer-2020-0267

6. Calabresi PA, Kieseier BC, Arnold DL, et al. Pegylated interferon beta-1a for relapsing-remitting multiple sclerosis (ADVANCE): a randomised, phase 3, double-blind study. The Lancet Neurology 2014;13(7):657–657. doi: 10.1016/S1474-4422(14)70068-7

7. Calabresi PA, Radue EW, Goodin D, et al. Safety and efficacy of fingolimod in patients with relapsing-remitting multiple sclerosis (FREEDOMS II): a double-blind, randomised, placebo-controlled, phase 3 trial. Lancet Neurol 2014;13(6):545–545. doi: 10.1016/s1474-4422(14)70049-3 [published Online First: 20140328]

8. Cohen JA, Barkhof F, Comi G, et al. Oral fingolimod or intramuscular interferon for relapsing multiple sclerosis. N Engl J Med 2010;362(5):402–402. doi: 10.1056/NEJMoa0907839 [published Online First: 20100120]

9. Cohen JA, Coles AJ, Arnold DL, et al. Alemtuzumab versus interferon beta 1a as first-line treatment for patients with relapsing-remitting multiple sclerosis: a randomised controlled phase 3 trial. Lancet 2012;380(9856):1819–1819. doi: 10.1016/s0140-6736(12)61769-3 [published Online First: 20121101]

10. Cohen JA, Comi G, Selmaj KW, et al. Safety and efficacy of ozanimod versus interferon beta-1a in relapsing multiple sclerosis (RADIANCE): a multicentre, randomised, 24-month, phase 3 trial. The Lancet Neurology 2019;18(11):1021–1021. doi: 10.1016/S1474-4422(19)30238-8

11. Coles AJ, Twyman CL, Arnold DL, et al. Alemtuzumab for patients with relapsing multiple sclerosis after disease-modifying therapy: a randomised controlled phase 3 trial. The Lancet 2012;380(9856):1829–1829. doi: 10.1016/S0140-6736(12)61768-1

12. Comi G, Kappos L, Selmaj KW, et al. Safety and efficacy of ozanimod versus interferon beta-1a in relapsing multiple sclerosis (SUNBEAM): a multicentre, randomised, minimum 12-month, phase 3 trial. Lancet Neurol 2019;18(11):1009–1009. doi: 10.1016/s1474-4422(19)30239-x [published Online First: 20190903]

13. Ebers GC. Randomised double-blind placebo-controlled study of interferon β-1a in relapsing/remitting multiple sclerosis. The Lancet 1998;352(9139):1498–1498. doi: 10.1016/S0140-6736(98)03334-0

14. Fox RJ, Miller DH, Phillips JT, et al. Placebo-controlled phase 3 study of oral BG-12 or glatiramer in multiple sclerosis. N Engl J Med 2012;367(12):1087–1087. doi: 10.1056/NEJMoa1206328

15. Giovannoni G, Comi G, Cook S, et al. A Placebo-Controlled Trial of Oral Cladribine for Relapsing Multiple Sclerosis. New England Journal of Medicine 2010;362(5):416–416. doi: 10.1056/nejmoa0902533

16. Gold R, Kappos L, Arnold DL, et al. Placebo-Controlled Phase 3 Study of Oral BG-12 for Relapsing Multiple Sclerosis. New England Journal of Medicine 2012;367(12):1098–1098. doi: 10.1056/NEJMoa1114287

17. Group TIMSS. Interferon beta-1b is effective in relapsing-remitting multiple sclerosis. I Clinical results of a multicenter, randomized, double-blind, placebo-controlled trial 1993;43(4):655–655. doi: 10.1212/wnl.43.4.655

18. Hartung H-P, Gonsette R, Konig N, et al. Mitoxantrone in progressive multiple sclerosis: a placebo-controlled, double-blind, randomised, multicentre trial. The Lancet 2002;360(9350):2018–2018. doi: 10.1016/S0140-6736(02)12023-X

19. Hauser SL, Bar-Or A, Cohen JA, et al. Ofatumumab versus Teriflunomide in Multiple Sclerosis. N Engl J Med 2020;383(6):546–546. doi: 10.1056/NEJMoa1917246

20. Hauser SL, Bar-Or A, Comi G, et al. Ocrelizumab versus Interferon Beta-1a in Relapsing Multiple Sclerosis. New England Journal of Medicine 2017;376(3):221–221. doi: 10.1056/nejmoa1601277

21. Jacobs LD, Cookfair DL, Rudick RA, et al. Intramuscular interferon beta-1a for disease progression in relapsing multiple sclerosis. The Multiple Sclerosis Collaborative Research Group (MSCRG). Ann Neurol 1996;39(3):285–285. doi: 10.1002/ana.410390304

22. Johnson KP, Brooks BR, Cohen JA, et al. Extended use of glatiramer acetate (Copaxone) is well tolerated and maintains its clinical effect on multiple sclerosis relapse rate and degree of disability. Copolymer 1 Multiple Sclerosis Study Group. Neurology 1998;50(3):701–701. doi: 10.1212/wnl.50.3.701

23. Kappos L, Bar-Or A, Cree BAC, et al. Siponimod versus placebo in secondary progressive multiple sclerosis (EXPAND): a double-blind, randomised, phase 3 study. The Lancet 2018;391(10127):1263–1263. doi: 10.1016/S0140-6736(18)30475-6

24. Kappos L, Fox RJ, Burcklen M, et al. Ponesimod Compared With Teriflunomide in Patients With Relapsing Multiple Sclerosis in the Active-Comparator Phase 3 OPTIMUM Study. JAMA Neurology 2021;78(5):558. doi: 10.1001/jamaneurol.2021.0405

25. Kappos L, Radue E-W, O’Connor P, et al. A Placebo-Controlled Trial of Oral Fingolimod in Relapsing Multiple Sclerosis. New England Journal of Medicine 2010;362(5):387–387. doi: 10.1056/nejmoa0909494

26. Naismith RT, Wolinsky JS, Wundes A, et al. Diroximel fumarate (DRF) in patients with relapsing–remitting multiple sclerosis: Interim safety and efficacy results from the phase 3 EVOLVE-MS-1 study. Multiple Sclerosis Journal 2020;26(13):1729–1729. doi: 10.1177/1352458519881761

27. O’Connor P, Wolinsky JS, Confavreux C, et al. Randomized trial of oral teriflunomide for relapsing multiple sclerosis. N Engl J Med 2011;365(14):1293–1293. doi: 10.1056/NEJMoa1014656

28. Polman CH, O’Connor PW, Havrdova E, et al. A Randomized, Placebo-Controlled Trial of Natalizumab for Relapsing Multiple Sclerosis. New England Journal of Medicine 2006;354(9):899–899. doi: 10.1056/nejmoa044397

29. Steinman L, Fox E, Hartung H-P, et al. Ublituximab versus Teriflunomide in Relapsing Multiple Sclerosis. New England Journal of Medicine 2022;387(8):704–704. doi: 10.1056/nejmoa2201904

30. Torkildsen Ø, Myhr K-M, Bø L. Disease-modifying treatments for multiple sclerosis – a review of approved medications. European Journal of Neurology 2016;23(S1):18–27. doi: 10.1111/ene.12883

31. Chedid T, Moisset X, Clavelou P. Rationale for off-label treatments use in primary progressive multiple sclerosis: A review of the literature. Revue Neurologique 2022;178(9):932–932. doi: 10.1016/j.neurol.2022.02.461

32. Munschauer FE, Kinkel RP. Managing side effects of interferon-beta in patients with relapsing-remitting multiple sclerosis. Clinical Therapeutics 1997;19(5):883–883. doi: 10.1016/S0149-2918(97)80042-2

33. Turner AP, Williams RM, Sloan AP, et al. Injection anxiety remains a long-term barrier to medication adherence in multiple sclerosis. Rehabil Psychol 2009;54(1):116–116. doi: 10.1037/a0014460

34. Jivraj F, Kang S, Reedie S, et al. The Patient and Clinician Assessment of Gastrointestinal (GI) Related Adverse Events Associated with Oral Disease-Modifying Therapies in Multiple Sclerosis: A Qualitative Study. Advances in Therapy 2022;39(11):5072–5072. doi: 10.1007/s12325-022-02250-x

35. Burks J, Marshall TS, Ye X. Adherence to disease-modifying therapies and its impact on relapse, health resource utilization, and costs among patients with multiple sclerosis. ClinicoEconomics and Outcomes Research 2017;9:251–60. doi: 10.2147/CEOR.S130334

36. Zhang Y, Salter A, Wallström E, et al. Evolution of clinical trials in multiple sclerosis. Therapeutic Advances in Neurological Disorders 2019;12:1756286419826547. doi: 10.1177/1756286419826547

